# Association between TM6SF2 rs58542926 T/C gene polymorphism and significant liver fibrosis: A meta-analysis

**DOI:** 10.1101/2020.10.11.20210690

**Authors:** Ting-Ting Mei, Jing Zhang, Shan Tang, Hai-Qing Guo, Xin-Huan Wei, Wen-Yan Zhang, Ya-Li Liu, Shan Liang, Zuo-Peng Fan, Li-Xia Ma, Li-Xia Qiu, Yi-Rong Liu, Hai-Bin Yu

**Author notes:** Correspondence Author: Hai-Bin Yu, Department Three of Liver Disease Center,Beijing YouAn Hospital, Capital Medical University, 8 Xitoutiao, Youanmenwai Street, Beijing, China,100069. Author contributions:Ting-Ting Mei and Zhang J contributed equally to this work. Ting-Ting Mei wrote the manuscript. Shan Tang, Guo HQ, and Wei XH searched and filtered the literature. Zhang WY, Liu YL, and Liang S selected and interpreted the data. Fan ZP, Ma LX, Liu YR, and Qiu LX revised the manuscript. Yu HB conceived the study. Yu HB was the corresponding author. In addition, every author read and approved the final manuscript. Conflict-of-interest statement:The authors have no conflict of interest to declare. There are no financial or other competing interests for principal investigators, the patients included, or any trial participant. Funding:This research did not receive any specific grant from funding agencies in the public, commercial, or not-for-profit sectors.

## Abstract

**Aim:** To further explore the association between Transmembrane 6 superfamily member 2 (TM6SF2) rs58542926 T/C gene polymorphism and hepatic fibrosis.

**Materials and Methods:** In this study the MEDLINE, PubMed, EMBASE, and CENTRAL databases were queried from inception to March 21, 2020. According to inclusion and exclusion criteria, case-control studies assessing the relationship between TM6SF2 rs58542926 T/C gene polymorphism and significant liver fibrosis were selected. NOS scale was used to evaluate the included literature. Stata 12.0 software was used for data analysis.

**Results:** In this meta-analysis,a total of 7 articles, including 2286 patients were included. Statistical analysis showed that the TM6SF2 gene polymorphism was associated with significant liver fibrosis in the allele contrast, recessive dominant models (T vs. C, OR=1.292, 95%CI 1.035-1.611, P=0.023; TT vs. CT+CC, OR=2.829, 95%CI 1.101-7.267, P=0.031). No significant publication bias was found after Egger’s test.

**Conclusion:** The present findings suggest that the TT genotype and T gene of TM6SF2 rs58542926 T/C gene polymorphism are associated with susceptibility to significant hepatic fibrosis.

## Introduction

Liver fibrosis is abnormal hyperplasia of the liver connective tissue caused by various pathogenic factors. Untreated liver fibrosis eventually leads to cirrhosis and is associated with an increased risk of developing hepatocellular carcinoma (HCC) ^[1,2]^. The most common causes of liver fibrosis are viral hepatitis C or B, alcohol abuse, schistosomiasis, and nonalcoholic fatty liver disease (NAFLD) ^[3]^. Yet, it still remains unclear whether the genetic factors aggravate the formation and development of liver fibrosis.

TM6SF2 is a gene located on chromosome 19, which encodes a segment of protein consisting of 351 amino acids ^[4]^. Yet, considering that the current data are contradictory, its function remains unknown. Yang-Lin *et al* confirmed that TM6SF2 is associated with histologically defined nonalcoholic fatty liver disease (NAFLD) and discovered that TM6SF2 serves as a powerful modifier of hepatic fibrogenesis and influences hepatic fibrosis progression in patients with non-alcoholic fatty liver disease ^[5]^. More recently, Zheng-Tao *et al* ^[6]^ conducted a meta-analysis and found that TM6SF2 rs58542926 T/C gene polymorphism was associated with the genetic susceptibility to hepatitis C virus (HCV) related hepatic fibrosis. However, Wong *et al* suggest that the TM6SF2 variant probably does not cause severe liver injury from NAFLD ^[7]^. In order to further clarify the relationship between TM6SF2 rs58542926 gene polymorphism and significant liver fibrosis, we conducted this meta-analysis.

## Materials and Methods

The current meta-analysis complied with Preferred Reporting Items for Systematic Reviews and Meta-Analyses (PRISMA) guidelines ^[8]^. The search strategy, eligibility criteria, and outcomes were described a priori (PROSPERO CRD42019128492).

### Data sources and search strategies

Medline, EMBASE, PubMed, and CENTRAL databases were comprehensively searched without language restriction for literature addressing the association of TM6SF2 rs58542926 T/C gene polymorphism with significant liver fibrosis published from inception to March 2020. The following key search terms and their potential combination were used “Cirrhosis, Liver OR Cirrhoses, Liver OR Hepatic Cirrhosis OR Cirrhoses, Hepatic OR Fibrosis, Liver OR Fibrosis, Liver” and “Transmembrane 6 superfamily member 2 OR TM6SF2”. The last literature search of the above databases was completed on March 21, 2020.

### Study selection

The relevant articles were initially selected by reading the title and abstract. Two authors reviewed the full texts to select qualified articles based on set eligibility criteria. Any disputes during the selection process were discussed and resolved by a third investigator.

### Inclusion and exclusion criteria

Inclusion criteria were the following: (1) the study cohorts included TM6SF2 rs58542926 T/C gene polymorphism in patients with significant liver fibrosis or cirrhosis and control individuals; (2) the included literature had clear criteria for staging of liver fibrosis or the diagnosis of cirrhosis; (3) case-control study and the control group included normal or non-significant liver fibrosis population; (4) if duplicate research reports of the same author or the same population were retrieved, then the complete data were selected for combined analysis to avoid duplicate statistics. (5) observation index: TM6SF2 rs58542926 T/C gene polymorphism frequency distribution; (6) the full text could be retrieved in different ways.

Exclusion criteria were: (1) the unclear source of enrolled cases in the article or lack of control group; (2) no clear criteria for the staging of liver fibrosis or the diagnosis of described cirrhosis; (3) unscientific or inappropriate data collection and analysis methods; (4) lack of detailed genotyping data; (5) no-case-control study; (6) animal research experiments; (7) literature that does not conform to the Hardy-Weinberg equilibrium (HWE).

### Assessment of evidence Quality

The Newcastle Ottawa Scale was used to assess the quality and risk of bias in case-control studies. This scale judges three general areas: a selection of study groups, comparability of groups, and the factor of exposure. We assessed the quality of evidence from the relevant studies for each outcome.

### Data extraction

Two experienced authors independently extracted the necessary data and information from eligible publications according to a predetermined data extraction form. The information extracted from all the selected studies included: first author’s surname, publication year, a country in which the study was conducted, age, body mass index (BMI), grading criteria for liver fibrosis or diagnostic criteria for cirrhosis, the numbers of cases and controls with the C/C, C/T, and T/T genotypes. Whether genotype distribution was consistent with the Hardy-Weinberg equilibrium (HWE) was also recorded.

### Risk of bias

Egger’s test was employed to assess publication bias. A P<0.05 was considered to be statistically significant.

### Statistical analysis

According to the requirements of Meta-analysis, a unified spreadsheet was designed to extract, sort, and transform the data, and the sorted data were rechecked with the original literature. Stata 12.0 software was used for data analysis. The association of the T/C polymorphism in the TM6SF2 gene with significant liver fibrosis was evaluated by calculating pooled odds ratios (ORs) alongside 95% confidence intervals (CIs) in the allelic, dominant, recessive, and super-dominant models. The HWE for each study was measured by the χ^2^ test, and *P*>0.05 was regarded as consistent with the HWE. The random or fixed-effects model was used to pool ORs based on heterogeneity assumption. Heterogeneity across studies was determined by the Q-and *I*^2^ tests. The fixed-effects model was used in the case of nonsignificant heterogeneity (*P*>0.05, *I*^2^<50%); otherwise, the random-effects model was utilized. In order to explore the effect of a single study on overall results, a sensitivity analysis was performed by removing one study sequentially to evaluate its effect on the overall results under all genetic models.

## Results

### Search results

There were 225 relevant studies compliant with the strategy, of which 68 were excluded as duplicates. After further review of the title and abstract, 65 reports were excluded as irrelevant to this meta-analysis. The second round of the evaluation was based on a careful full-text review of the 92 retained papers. Then, 85 reports were excluded, leaving seven studies that were included in the final analysis. **Figure** Supplementary **1** summarizes the above selection process.

### Characteristics of the included studies

**Table 1** describes the basic features of all seven included studies. A total of 719 patients with significant liver fibrosis (case group) and 1567 patients without significant liver fibrosis (control group) were included. Genotyping data for all studies are summarized in **Table 2**. The vast majority of reports used TaqMan assays for genotyping. The assessed individuals were mostly Europeans and Asians.

**Table 1.**
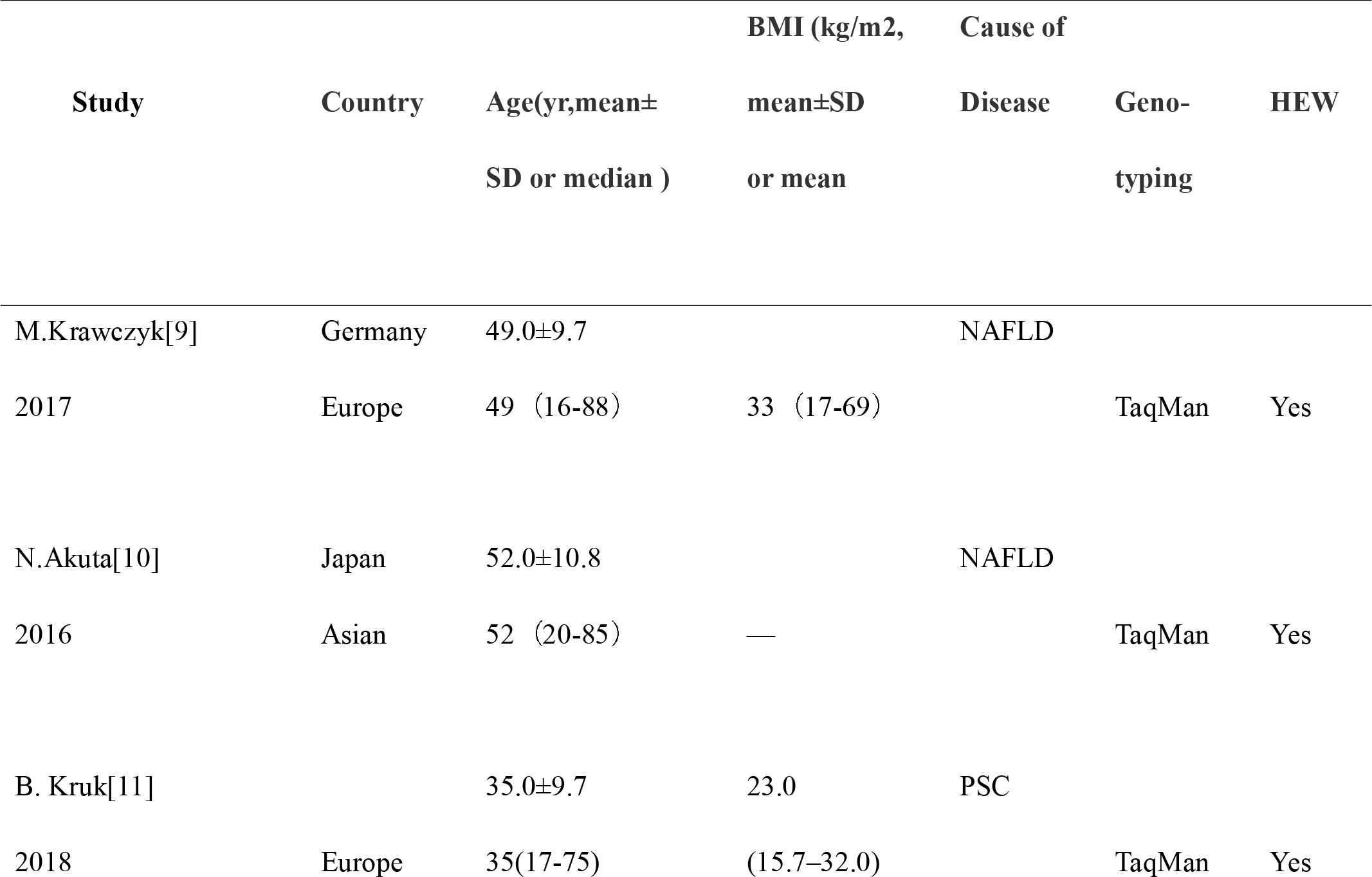
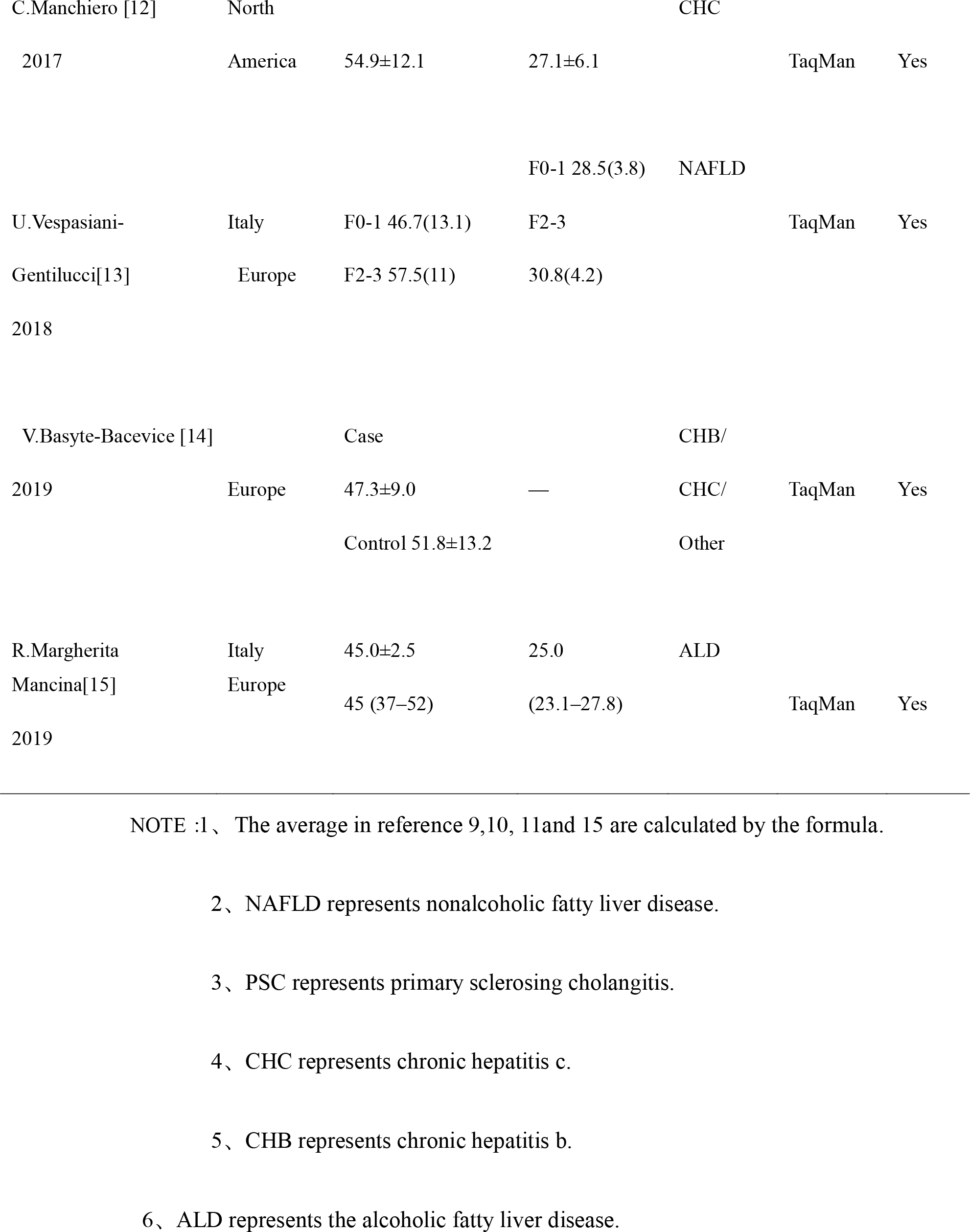
Characteristics of the studies included in the meta-analysis

**Table 2.** Diagnosis and grading of liver fibrosis and the distribution of the TM6SF2 genotypes in patients

| Study | Method of grading or diagnosis | Genotype<br>(CC/CT/TT ) | Fibrosis<br>grading or<br>cirrhosis<br>( Control ) | Fibrosis<br>grading or<br>cirrhosis<br>( Case ) |
| --- | --- | --- | --- | --- |
| M.Krawczyk<sup>[9]</sup> |  | case 68/20/2 |  |  |
| 2017 | Kleiner Score | control 164/38/3 | 0-1 | 2-3 |
| N.Akuta<sup>[10]</sup> |  | case 41/13/1 |  |  |
| 2016 | Brunt Score | control 63/20/1 | 0-2 | 3-4 |
| B. Kruk<sup>[11]</sup> | Diagnostic criteria for cirrhosis | case 50/5/0 |  |  |
| 2018 |  | control 107/16/0 | No | YES |
| C.Manchiero<sup>[12]</sup> | Metavir Score | case 46/12/0 |  |  |
| 2017 |  | control 215/17/0 | 0-2 | 3-4 |
| U.Vespasiani-Gentilucci<sup>[13]</sup> | Kleiner Score | case 32/8/2 |  |  |
| 2018 |  | control 42/9/0 | 0-1 | 2-3 |
| V.Basyte-Bacevicius<sup>[14]</sup> | Diagnostic criteria for cirrhosis | case 279/53/2 |  |  |
| 2019 |  | control 471/77/2 | No | YES |
| R.Margherita<br>Mancina<sup>[15]</sup><br>2019 | Diagnostic criteria<br>for cirrhosis | case71/11/3<br>control 277/44/1 | No | YES |

NOTE: Diagnosis of liver cirrhosis was established by standard clinical features, laboratory tests, and radiological imaging.

### Literature quality evaluation

The 7 included studies were scored with NOS. The results showed that all the literature was rated with at least 7, which indicated that the seven kinds of literature were of high quality (**Table 3**).

**TABlE3.**
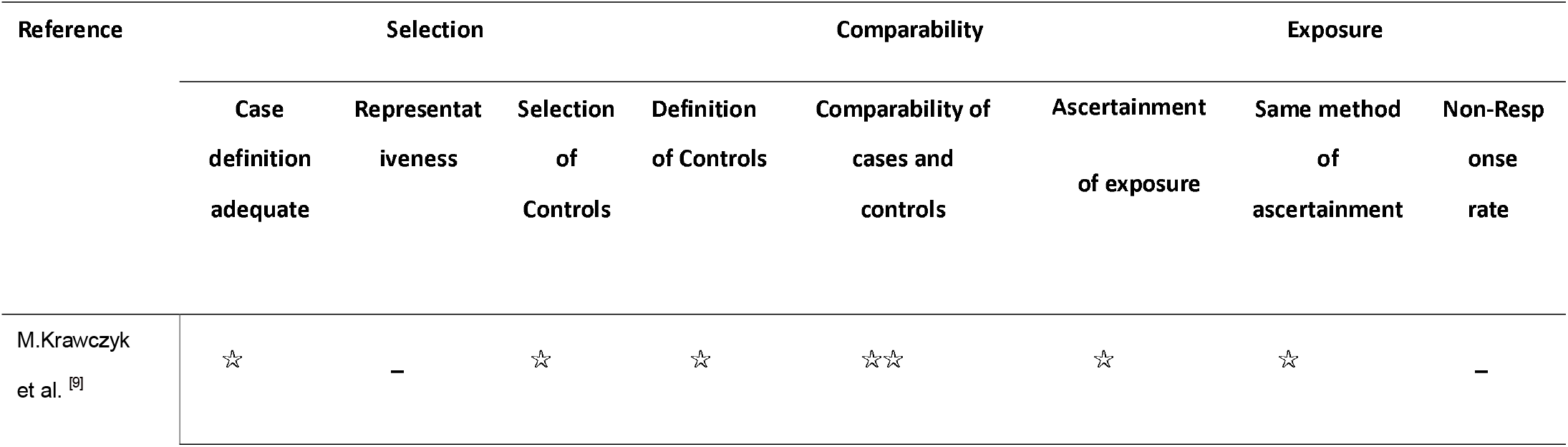
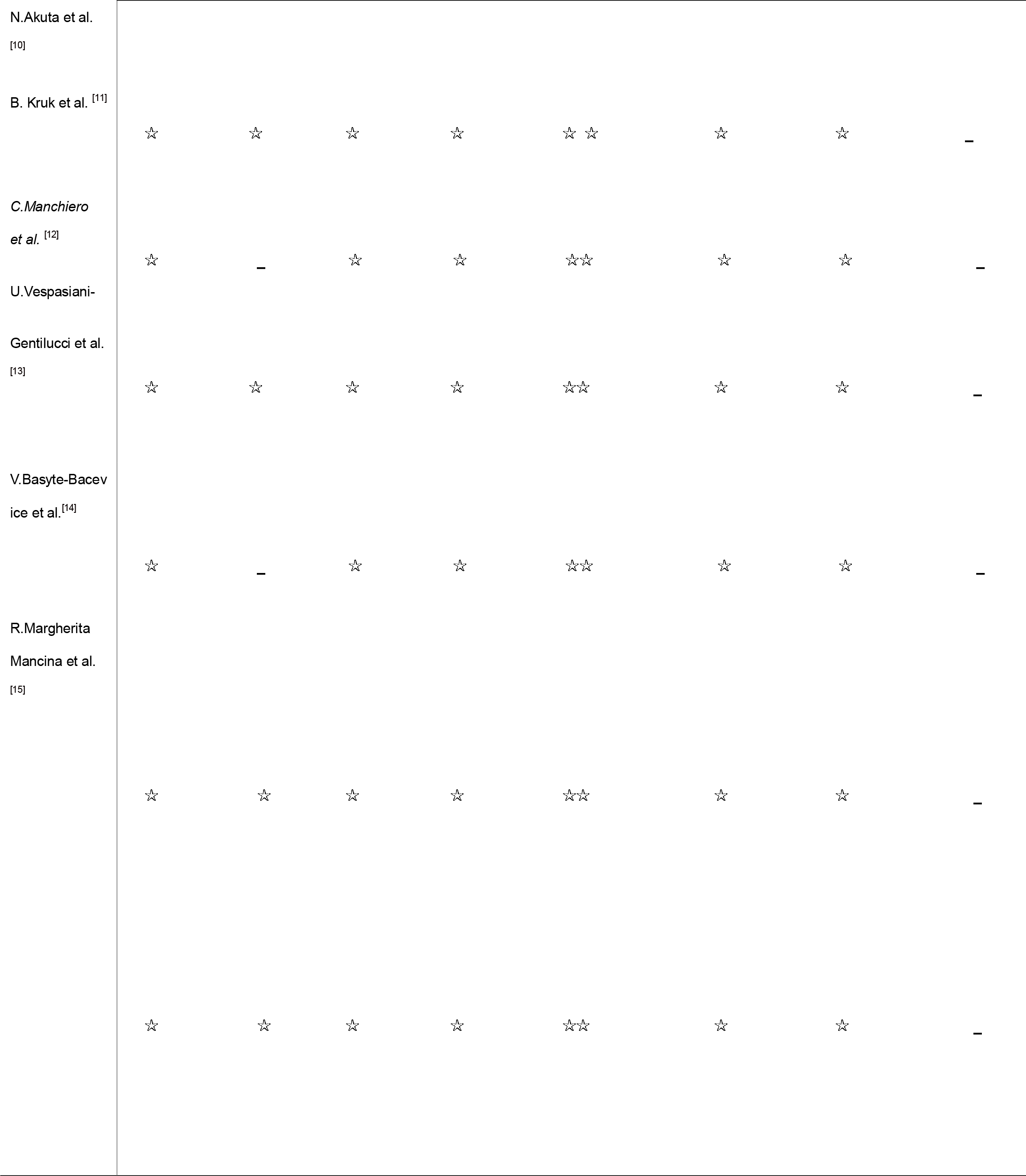
Summary of critical appraisal of included studies using the Newcastle–Ottawa Quality Assessment Scale for case-control studies.

### Meta-analysis results

Seven studies included in the current meta-analysis described the association of TM6SF2 rs58542926 T/C gene polymorphism with significant liver fibrosis. The allelic (T vs. C), dominant (CT+TT vs. CC), recessive (TT vs. CT+CC), and super-dominant (CC+TT vs. TC) models were assessed. The fixed-effects model was employed for pooled ORs since nonsignificant heterogeneities were detected in allelic, dominant, and recessive models. In the super-dominant model, heterogeneity was significant, treatment: (1) conducted sensitivity analysis found that the results of this study were robust, (2) subgroup analysis was performed according to age and sample size. The results showed that age and sample size were not the source of heterogeneity (**Table S1**). Therefore, the random effect model was employed for pooled ORs. The sources of heterogeneities were considered as follows: (1) the source of each study population was not completely the same, and there were few data on population in some areas, this subgroup analysis was not performed, considering that different populations may be the source of heterogeneity; (2) the BMI and grading criteria of hepatic fibrosis or diagnostic criteria of cirrhosis distribution of each study population were not the same, which may also be the source of heterogeneity. This meta-analysis found that the TT genotype and T gene of TM6SF2 E167K gene polymorphism were associated with susceptibility to significant hepatic fibrosis.

### TM6SF2 rs58542926 T/C in the dominant model (CT+TT vs. CC)

The CT+TT genotype as the exposure factor and the CC genotype as the non-exposure factor were analyzed. A total of 132 and 587 cases had the TT+CT and CC genotypes in the case group, respectively. Meanwhile, 228 and 1339 cases had the TT+CT and CC genotypes in the control group, respectively. The results showed that the pooled risk of significant liver fibrosis was not significantly different in the TT+CT genotype compared with the CC genotype (CT+TT vs. CC, OR=1.257; 95%CI 0.990-1.595; P=0.060; **Figure 1**).

**Fig. 1.**
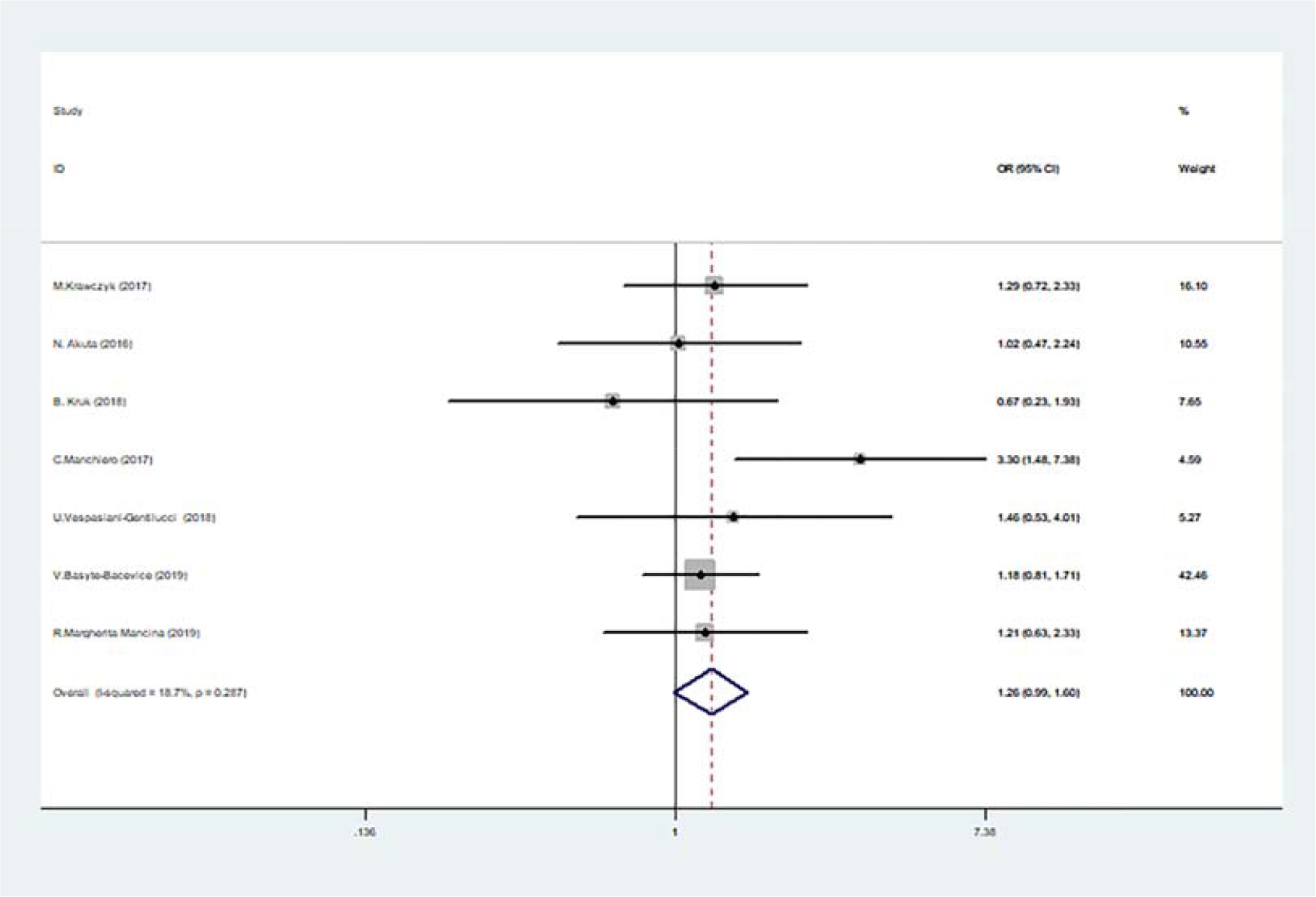
Forest plot of studies evaluating the OR with 95%CI of TM6SF2 rs58542926 T/C in the dominant model (CT□+ □ TT vs. CC) in significant liver fibrosis patients. CI, Confidence interval; OR, odds ratio

### TM6SF2 rs58542926 T/C in the allelic model (T vs. C)

The T allele was used as the exposure factor and the C allele as the non-exposure factor. There were 142 cases with the T allele and 1296 with the C allele in the case group, and 235 T allele and 2899 C allele cases in the control group. We found that TM6SF2 rs58542926 T/C gene polymorphism had a significant association with significant liver fibrosis. (T vs C, OR=1.292; 95%CI 1.035-1.611; P=0.023; **Figure 2**)

**Fig. 2.**
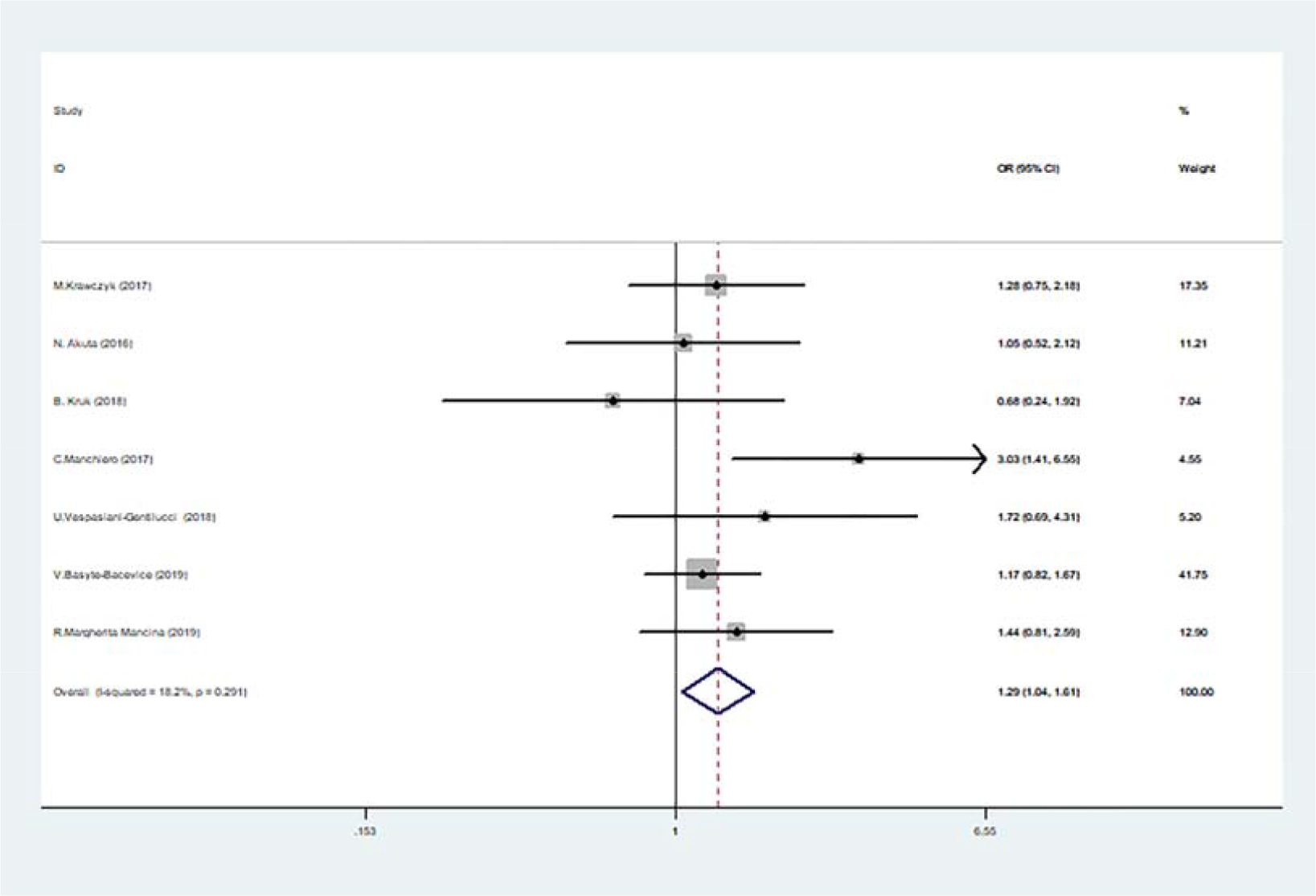
Forest plots of studies evaluating the OR with 95%CI of TM6SF2 rs58542926 T/C in the allelic model (T vs. C) in significant liver fibrosis patients. CI, Confidence interval; OR, odds ratio

### TM6SF2 rs58542926 T/C in the recessive model (CT+CC vs TT)

The TT genotype was used as the exposure factor and the CC+CT genotype as the non-exposure factor. Ten patients had the TT genotype, and 709 displayed the CC+CT genotype among cases. Meanwhile, 7 and 1560 cases had the TT and CC+CT genotypes among controls, respectively. The results showed that the risk of significant liver fibrosis in the TT genotype group was higher than that of the CC+CT genotype group (TT vs. CT+CC, OR=2.829; 95%CI 1.101-7.267; P=0.031; **Figure 3**).

**Fig. 3.**
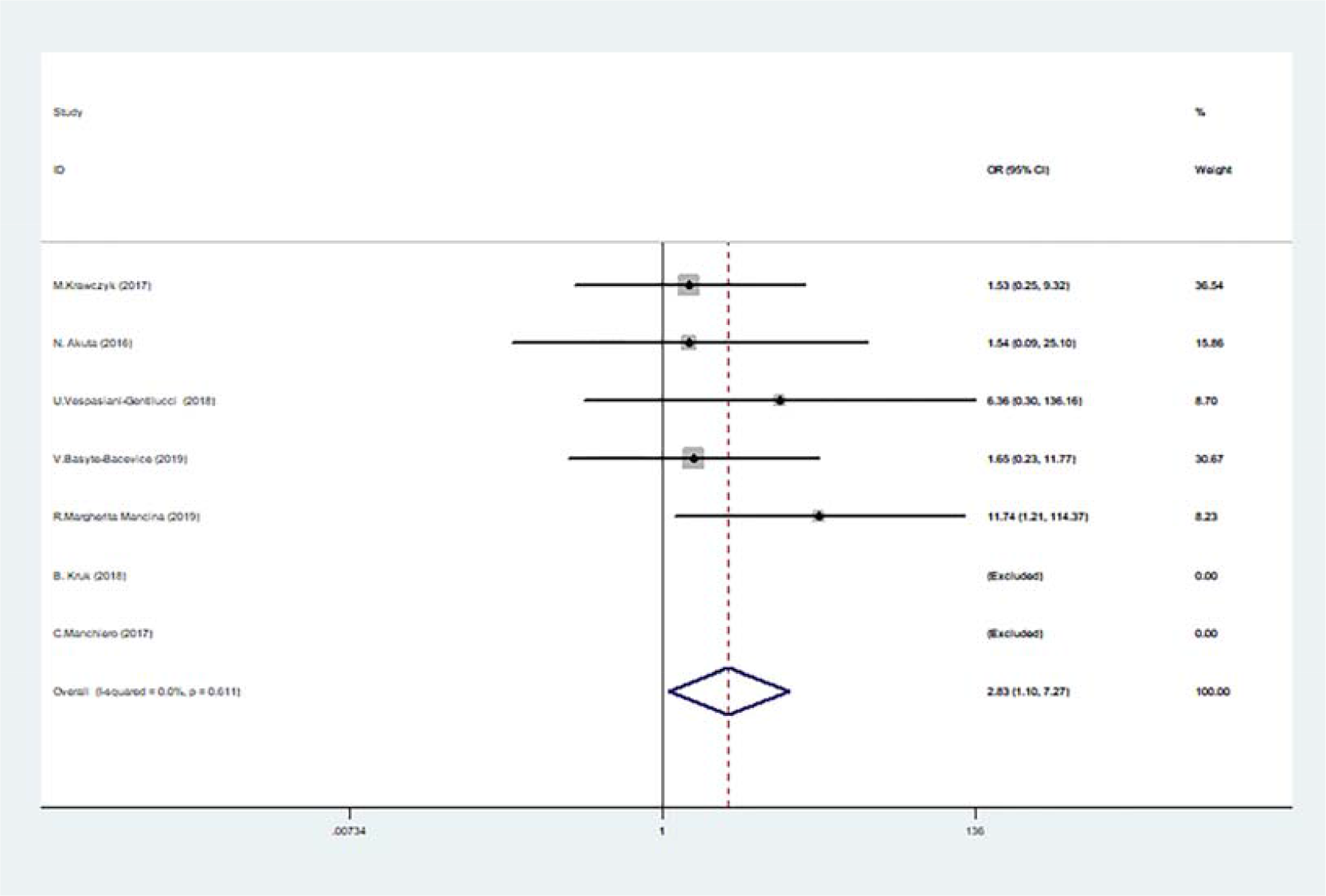
Forest plot of studies evaluating the OR with 95%CI of TM6SF2 rs58542926 T/C in the recessive model (CC□+ □CT vs. TT) in significant liver fibrosis patients. CI, Confidence interval; OR, odds ratio

### TM6SF2 rs58542926 T/C in the super-dominant model (CC+TT vs TC)

The TT+CC genotype was used as the exposure factor and the CT genotype as the non-exposure factor. There were 597 cases with the TT+CC genotype and 122 with the CT genotype in the case group. Meanwhile, 1302 and 265 cases had the TT+CC and CT genotypes in the control group, respectively. The results showed that the pooled risk of significant liver fibrosis was not significantly different in the TT+CC genotype compared with the CT genotype (CC+TT vs TC, OR=1.163; 95%CI 0.569-2.379; P=0.678; **Figure 4**).

**Fig. 4.**
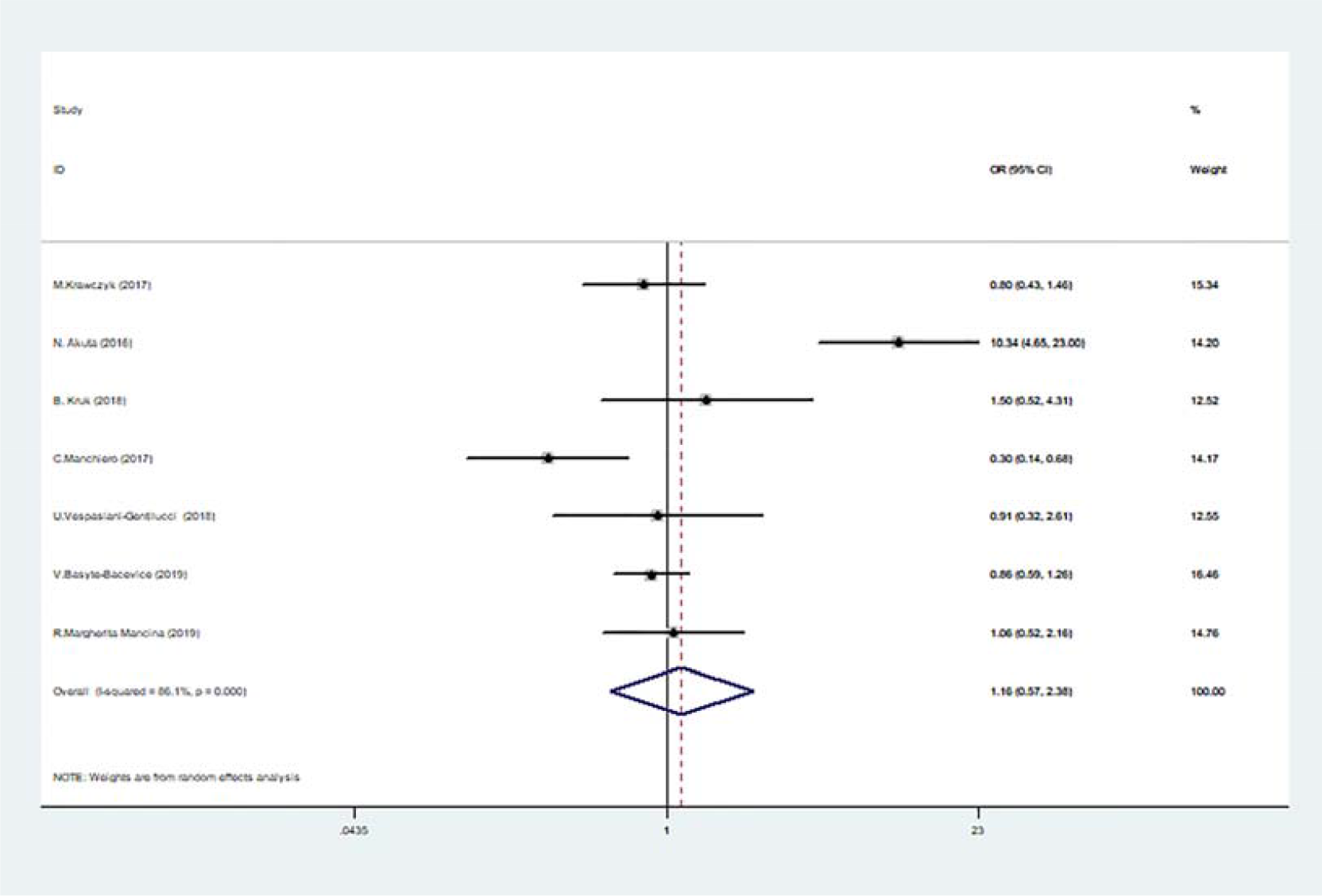
Forest plot of studies evaluating the OR with 95%CI of TM6SF2 rs58542926 T/C in the super-dominant model (CC□+ □TT vs. TC) in significant liver fibrosis patients. CI, Confidence interval; OR, odds ratio

### Sub-analysis

To analyze the source of heterogeneity in the super-dominant model, we performed subgroup analysis.

The results of subgroup analysis according to sample size in the super-dominant model (CC+TT vs. TC) suggested no interaction between sample size >200 groups and sample size ≤200 group (OR=1.163, 95% CI0.569-2.379, p=0.678). The sample size was not the source of heterogeneity (**Figure S2**).

Subgroup analyses, according to age in the super-dominant model (CC+TT vs. TC), were also conducted. The 95% CI of the two subgroups (Mean age (yr) >50 vs. Mean age (yr) ≤50) overlapped, the p-value of the subgroups was greater than 0.05 (OR=1.309, 95% CI 0.430-3.983, p=0.636). So, it was considered that there was no interaction between the subgroups, i.e., age was not the source of heterogeneity (**Figure S3**).

### Sensitivity analysis

Sensitivity analysis was performed by sequentially omitting one study so as to examine its effect on the overall results under all genetic models. In the four genetic models of TM6SF2 rs58542926 T/C, OR values obtained after eliminating any one of the studies were close to pre-exclusion ORs, indicating the robustness of the current analysis (**Figures S4-7**).

### Publication bias

Egger’s funnel plots showed no publication bias in the four genetic models, including the dominant (CT+TT vs. CC, P=0.267), allelic (T vs. C, P=0.497), recessive (TT vs. CT+CC, P=0.582) and super-dominant (CC+TT vs. TC, P=0.540) models (**Figures S8-11**).

## Discussion

According to the subcellular localization analysis, TM6SF2 is mainly expressed in the endoplasmic reticulum-Golgi body intermediate ^[4]^ and is polymorphic. The polymorphism of TM6SF2 rs58542926 refers to the substitution of thymine T for normal cytosine C in the nucleotide no. 499 of this gene, resulting in the substitution of glutamic acid (Glu) at no. 167 by lysine (Lys). This amino acid substitution of TM6SF2 rs58542926 (glutamate negatively charged and lysine positively charged) produces an unstable protein that reduces the secretion of very-low-density lipoprotein-mediated neutral fats (triglycerides and cholesterol esters) from hepatocytes, resulting in lipid droplet accumulation and increased levels of triglyceride in liver cells, which, in turn, cause severe fibrosis or cirrhosis ^[16, 17]^.

Although studies on the relationship between TM6SF2 rs58542926 T/C gene polymorphism and the risk of liver fibrosis and cirrhosis have attracted the attention of many researchers, different studies have reported contradictory results. Since single studies tend to have a small sample size, this might affect the stability and reliability of reported research results. A meta-analysis is a valid, scientific, and objective way to evaluate and combine results from different studies.

In this study, through the formulation of search strategies, data screening, and data extraction, seven documents that met the requirements were finally included in the meta-analysis. The data of TM6SF2 rs58542926 T/C gene polymorphism and significant liver fibrosis at different locations and at different times were combined and analyzed. We then evaluated whether TM6SF2 rs58542926 T/C gene polymorphism was associated with significant liver fibrosis and what was the strength of the correlation. The TM6SF2 E167K gene was analyzed for a dominant gene model, an allele model, a recessive gene model, and a super dominant gene model. The results suggested that the TT genotype and T gene of TM6SF2 rs58542926 T/C gene polymorphism were associated with susceptibility to significant hepatic fibrosis. This is consistent with the results of a meta-analysis conducted by Zheng-Tao *et al* ^[6]^ who found that TM6SF2 rs58542926 T/C gene polymorphism was associated with the genetic susceptibility to HCV related hepatic fibrosis, and the T allele at 499 was positively correlated with the susceptibility to hepatic fibrosis, suggesting that this allele may be a risk factor for hepatic fibrosis in patients with chronic hepatitis C. However, Zheng-Tao and his team only studied the relationship between this gene polymorphism and hepatitis c-related liver fibrosis, but did not explain the association between other causes of liver fibrosis and this gene polymorphism. Besides, they only investigated the recessive gene model. In this meta-analysis, we further explored the relationship between TM6SF2 rs58542926 T/C gene polymorphism and significant liver fibrosis, including HCV and other reasons causing significant liver fibrosis. In addition, more gene models were analyzed (recessive gene, a dominant gene, gene, super dominant genetic model); thus, making the results more representative. The inconsistent results may be explained by a variety of reasons. A meta-analysis can search for evidence as possible to analyze this difference and solve the controversial issue. But, a meta-analysis must not subjectively exclude research, and try to control and analyze various biases in order to improve the authenticity and reliability of meta-analysis results. This study was conducted in accordance with the requirements of the Newcastle Ottawa Scale to evaluate the quality of the literature, excluding uncontrolled, unclear diagnostic criteria, and duplicate reports. Publication bias is one of the critical factors influencing the results of a meta-analysis. The Egger linear regression results of this study suggested that there was no obvious publication bias in the literature included in this meta-analysis.

The study has some limitations. Firstly, single-factor research was involved, and the interaction of TM6SF2 E167K gene polymorphism with environmental factors and multi-gene linkage were not included; the interaction of environmental factors and multi-gene linkage can affect the susceptibility to liver fibrosis and cirrhosis. Secondly, due to the limited number of articles included, this meta-analysis failed to cause of disease and BMI for group discussion. Thirdly, the classification criteria for liver fibrosis included in this study differed. In addition, there may be slight differences in the definition of significant liver fibrosis. The result of subgroup analysis, according to fibrosis criteria, was not reliable because of few papers included in this meta (**Table S1**). Finally, the included literature mainly analyzed the relationship between TM6SF2 E167K gene polymorphism in the European population and significant liver fibrosis and cirrhosis. There are relatively few related studies from other countries and races. Therefore, this meta-analysis cannot be generalizable to all regions or ethnicities. The analysis also makes it impossible to compare the effect of this gene polymorphism on liver fibrosis in different ethnic populations.

## Conclusion

This meta-analysis suggests that the TT genotype and T gene of TM6SF2 rs58542926 T/C gene polymorphism are associated with susceptibility to significant hepatic fibrosis.

## Data Availability

All the data mentioned in the manuscript are available.

## Abbreviations

TM6SF2: transmembrane 6 superfamily member 2
HCC: hepatocellular carcinoma
NAFLD: nonalcoholic fatty liver disease
HCV: hepatitis C virus
PSC: primary sclerosing cholangitis
CHC: chronic hepatitis c
CHB: chronic hepatitis b
ALD: alcoholic fatty liver disease

## Acknowledgments

Not applicable

## Supplementary materials

**Fig. S1.**
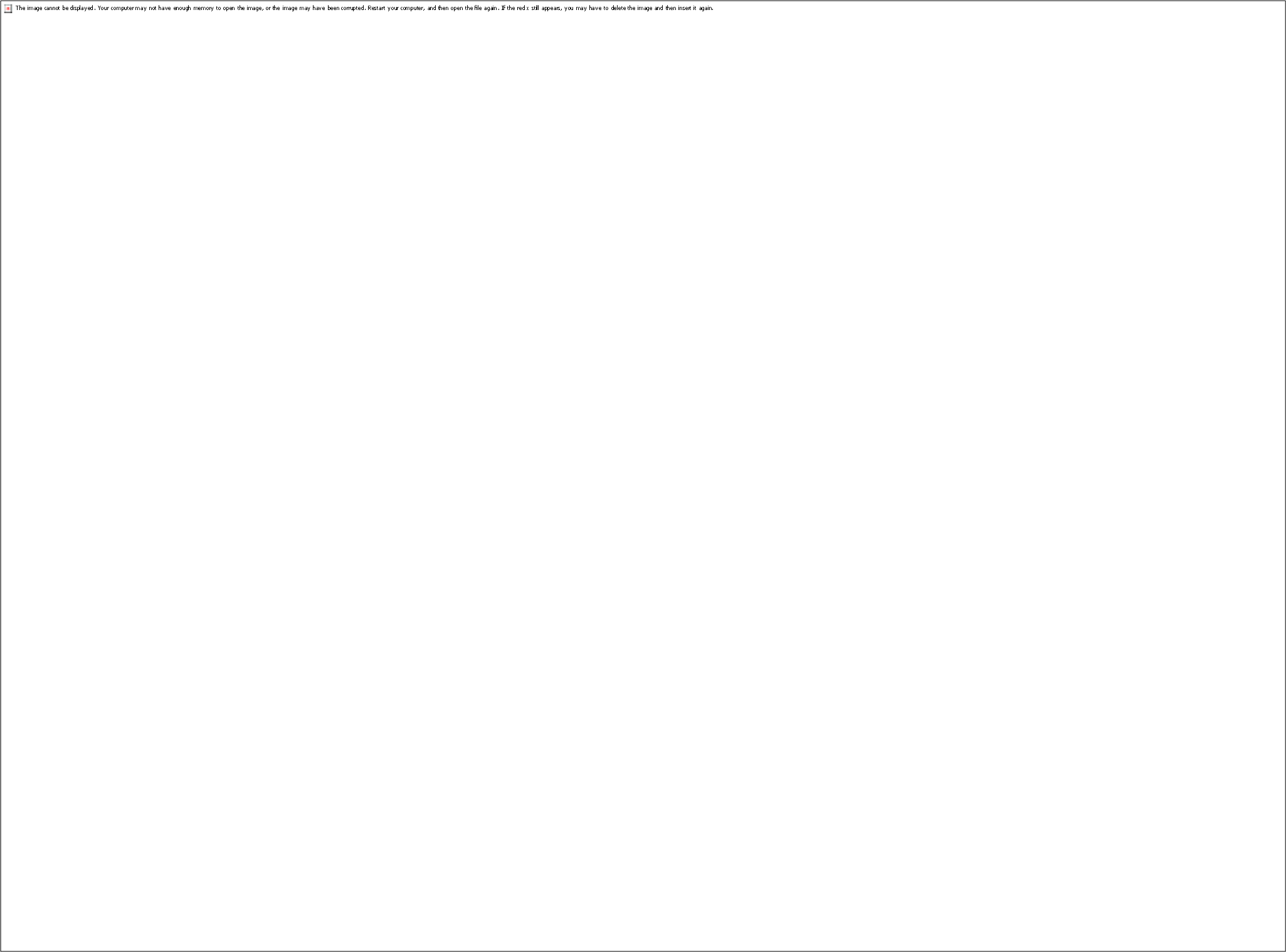
Forest plot of studies evaluating the OR with 95%CI of TM6SF2 rs58542926 T/C in the super-dominant model (CC□+ □TT vs. TC) in the subgroup analysis of sample size. CI, Confidence interval; OR, odds ratio.

**Fig. S2.**
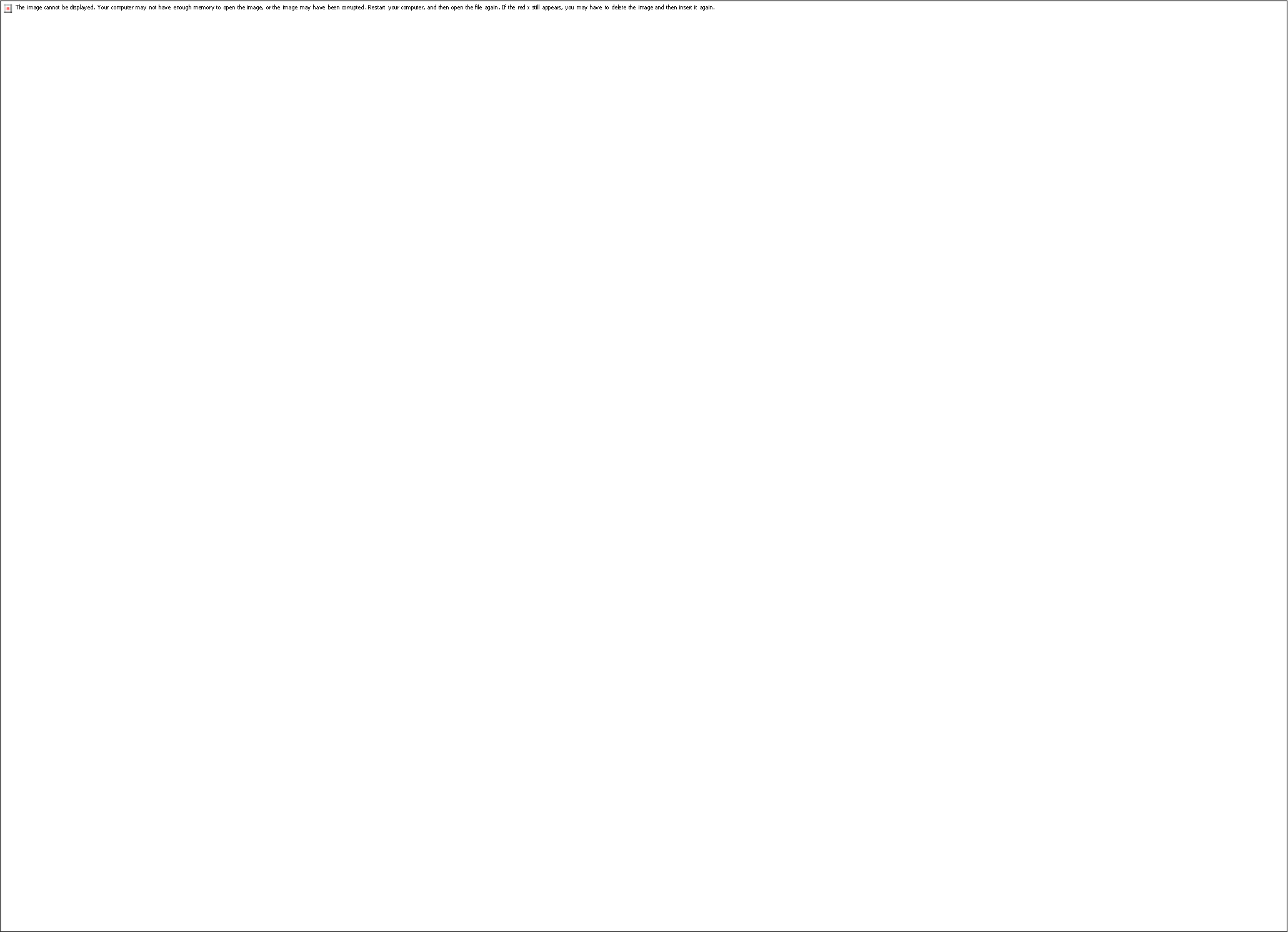
Forest plot of studies evaluating the OR with 95%CI of TM6SF2 rs58542926 T/C in the super-dominant model (CC□+ □TT vs. TC) in the subgroup analysis of patients with different Mean age(yr). CI, Confidence interval; OR, odds ratio.

**Fig. S3.**
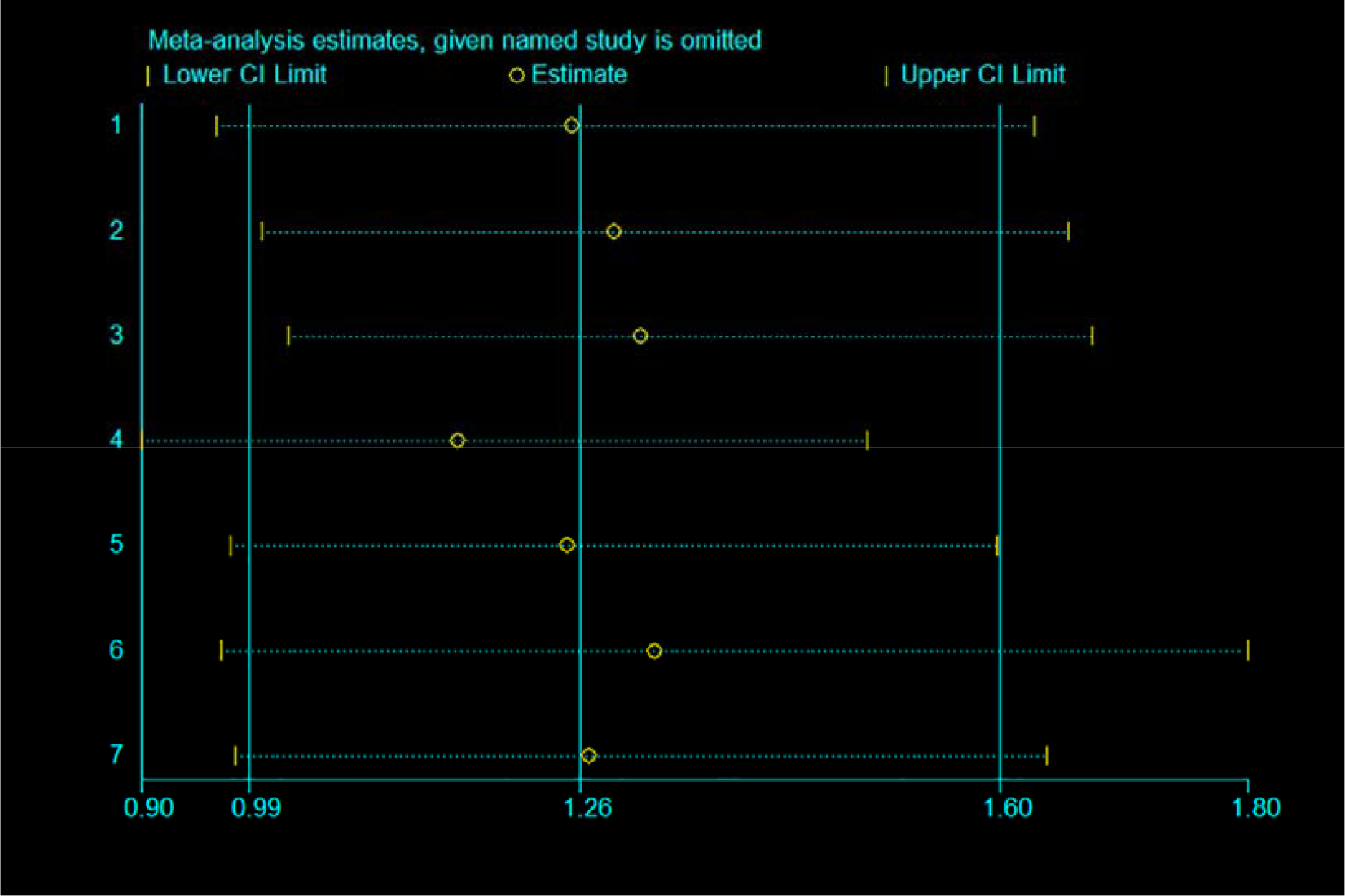
Sensitivity analysis of TM6SF2 rs58542926 T/C in the dominant model (CT□+ □TT vs. CC).

**Fig. S4.**
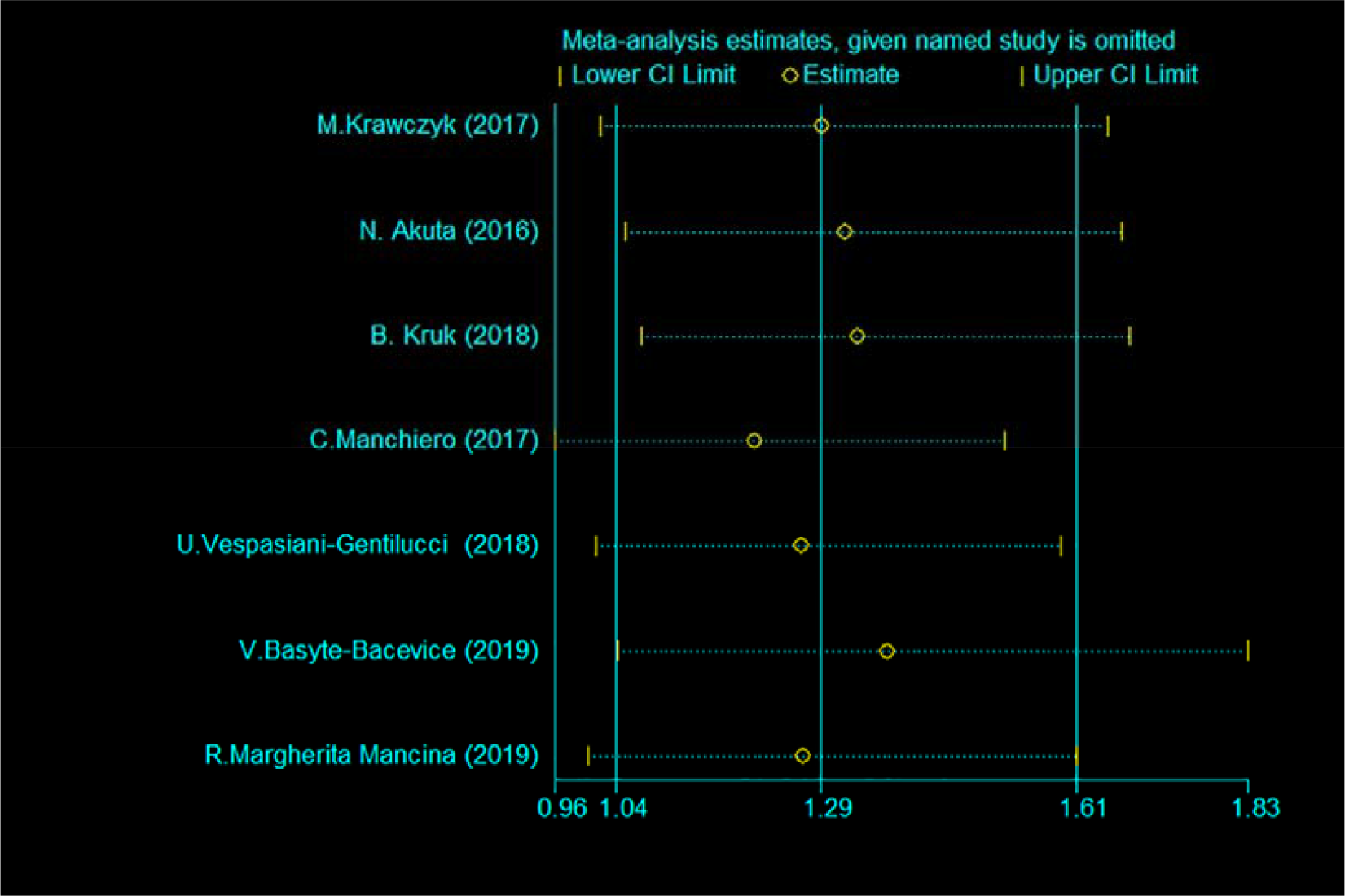
Sensitivity analysis of TM6SF2 rs58542926 T/C in the allelic model (T vs. C).

**Fig. S5.**
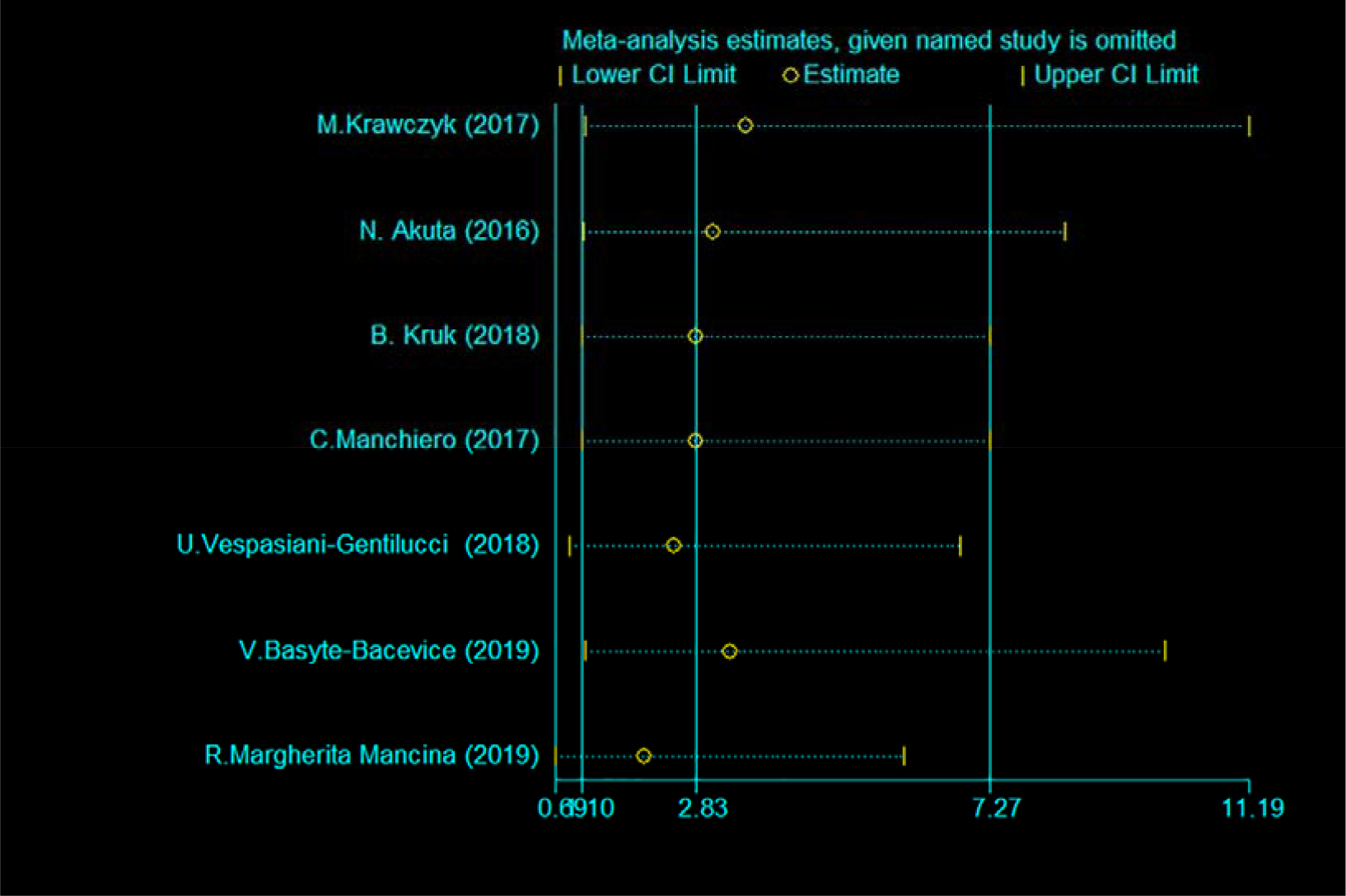
Sensitivity analysis of TM6SF2 rs58542926 T/C in the recessive model (TT vs. TC+CC).

**Fig. S6.**
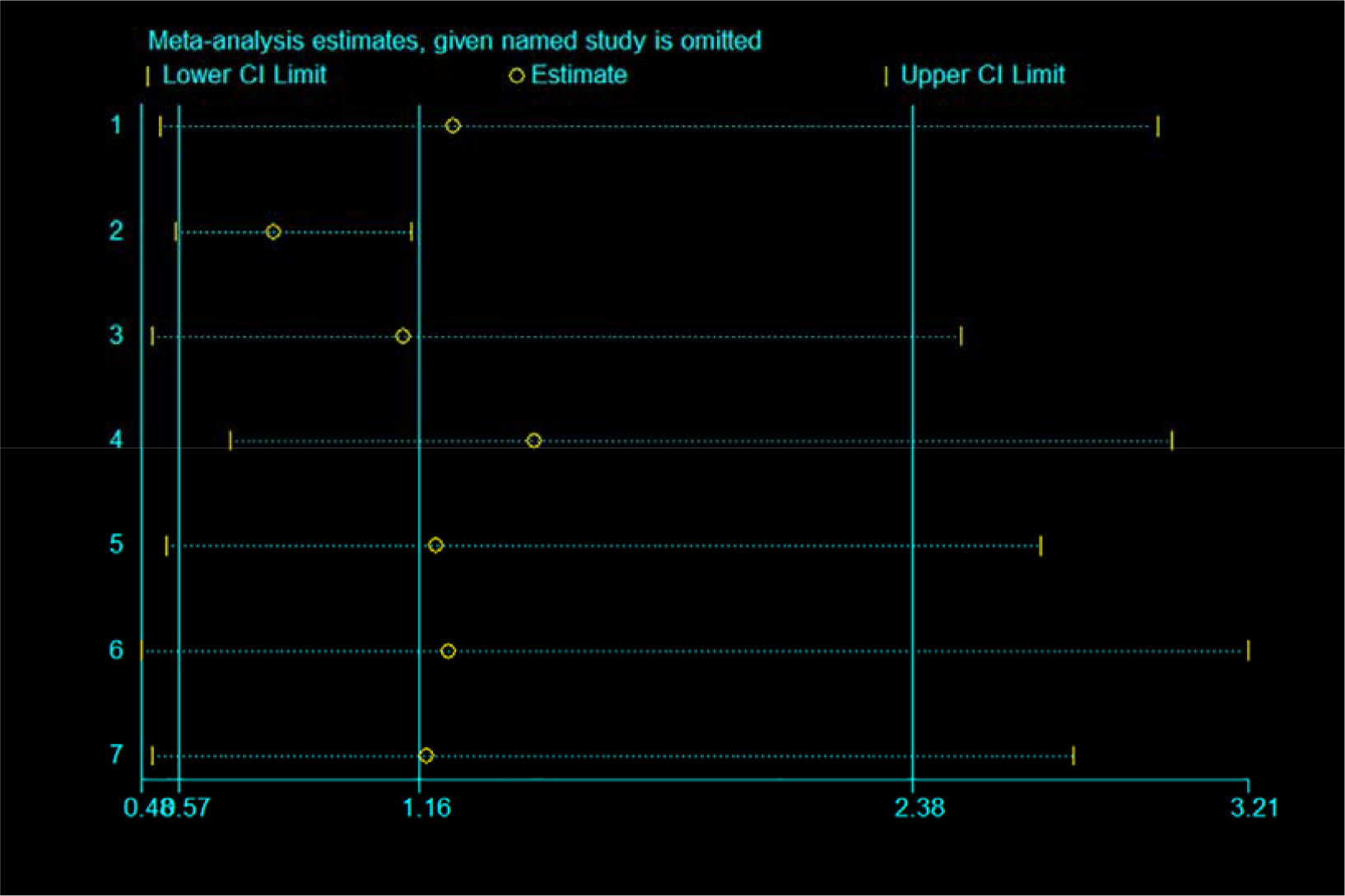
Sensitivity analysis of TM6SF2 rs58542926 T/C in the super-dominant model (TT+CCvs. TC).

**Fig. S7.**
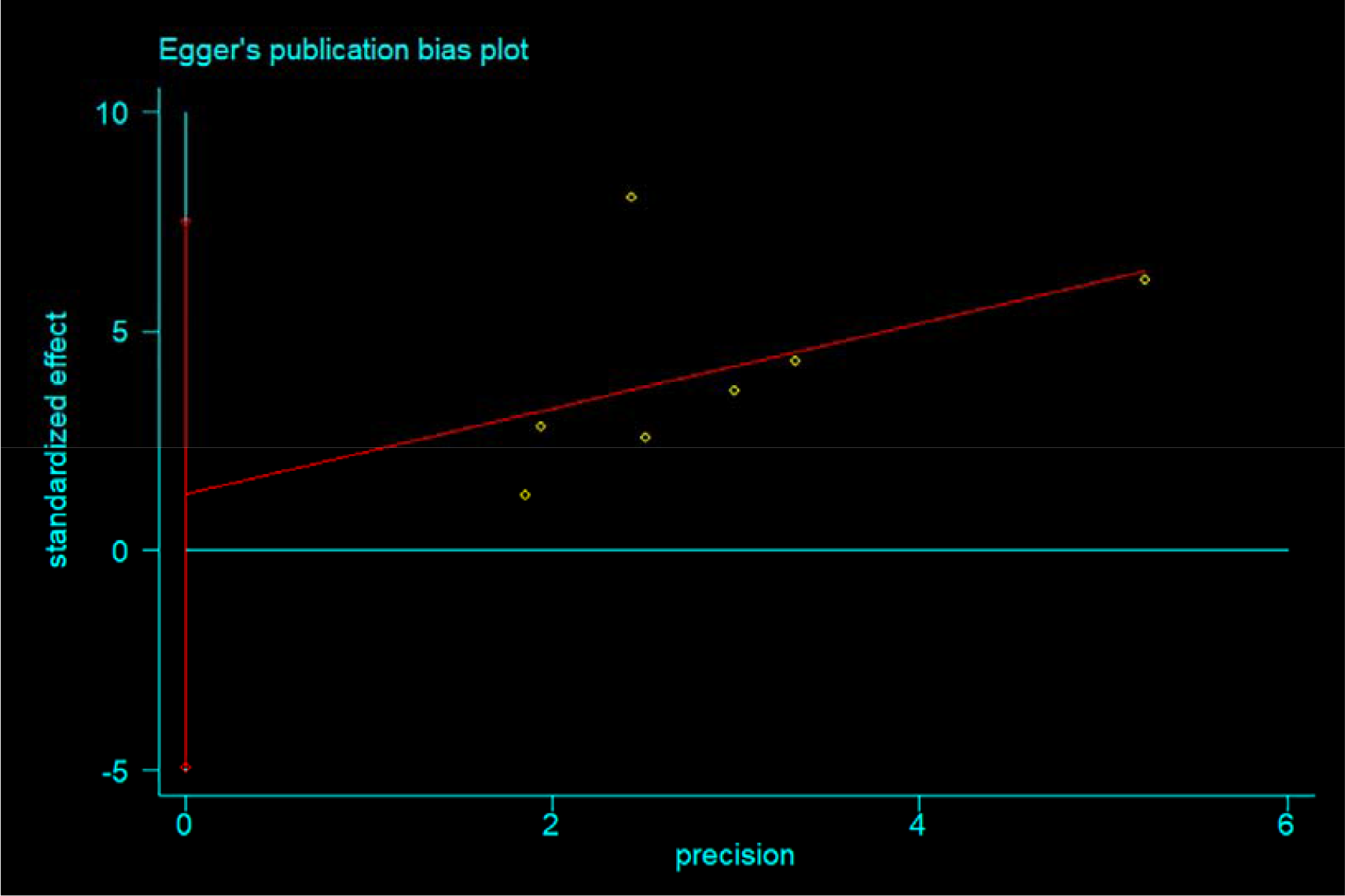
Egger’s funnel plot of TM6SF2 rs58542926 T/C in the dominant model (CT+TT vs. CC, P=0.267)

**Fig. S8.**
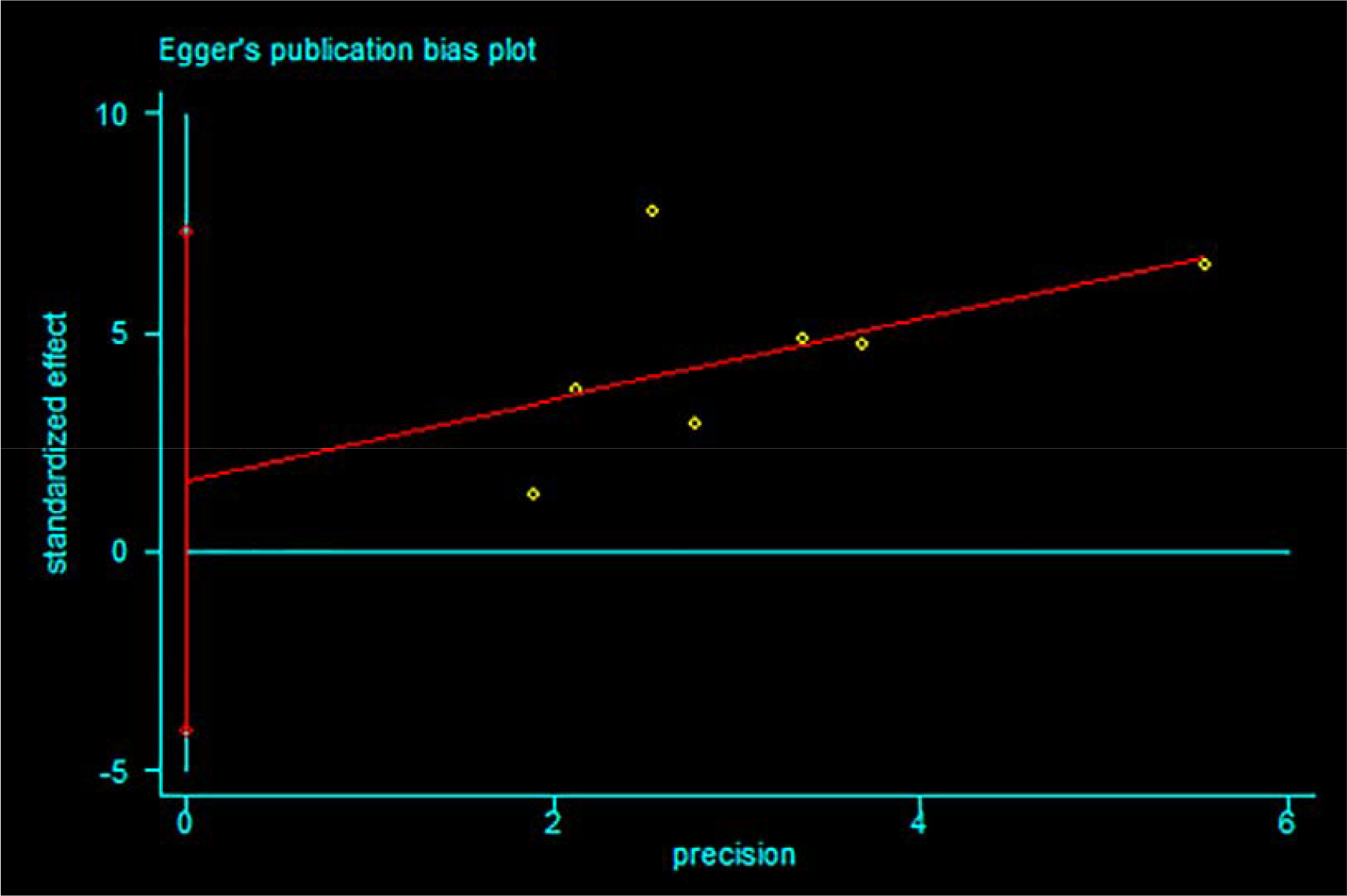
Egger’s funnel plot of TM6SF2 rs58542926 T/C in the allelic model (T vs C, P=0.497)

**Fig. S9.**
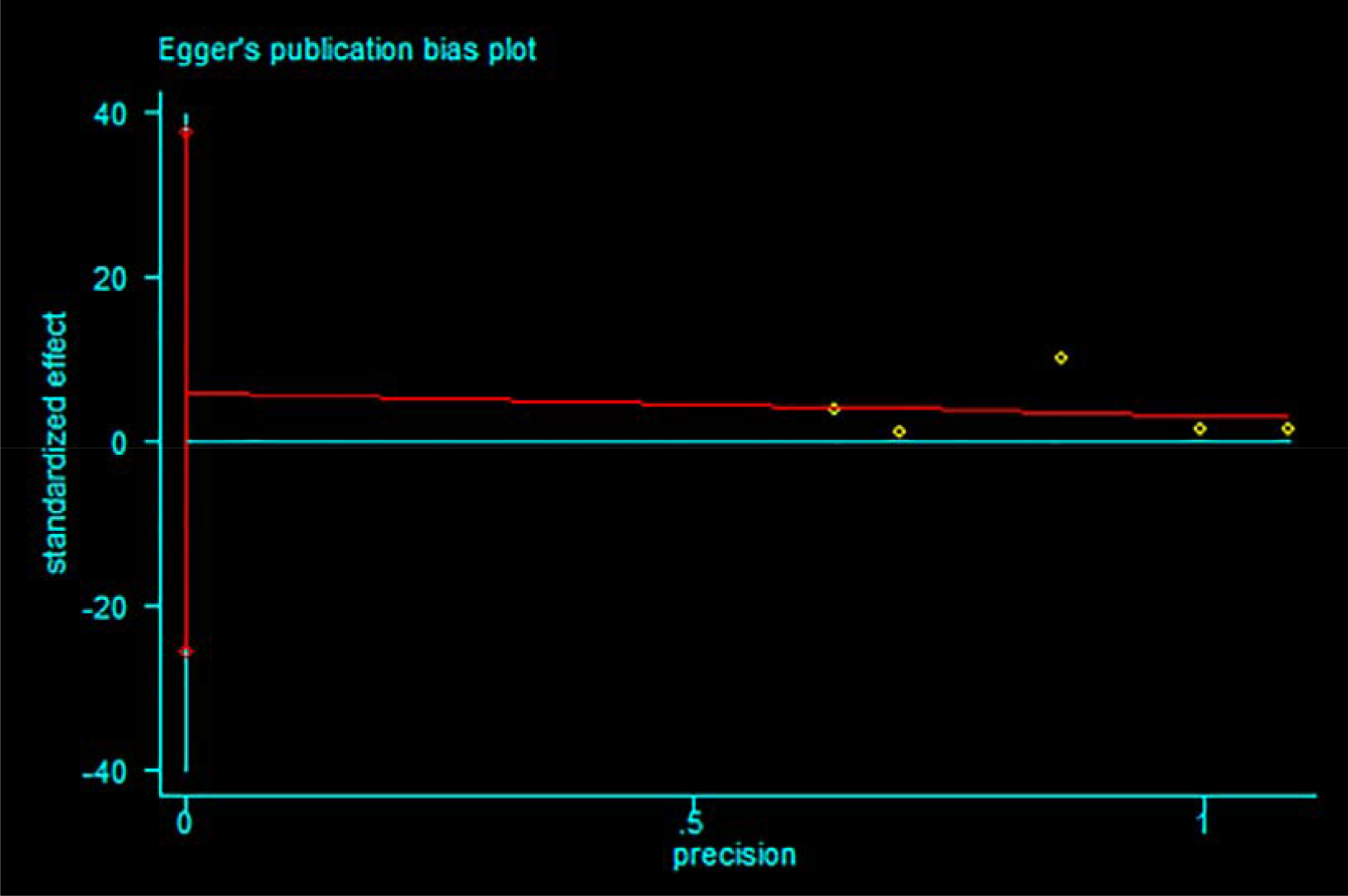
Egger’s funnel plot of TM6SF2 rs58542926 T/C in the recessive model (TT vs. CT+CC, P=0.582)

**Fig. S10.**
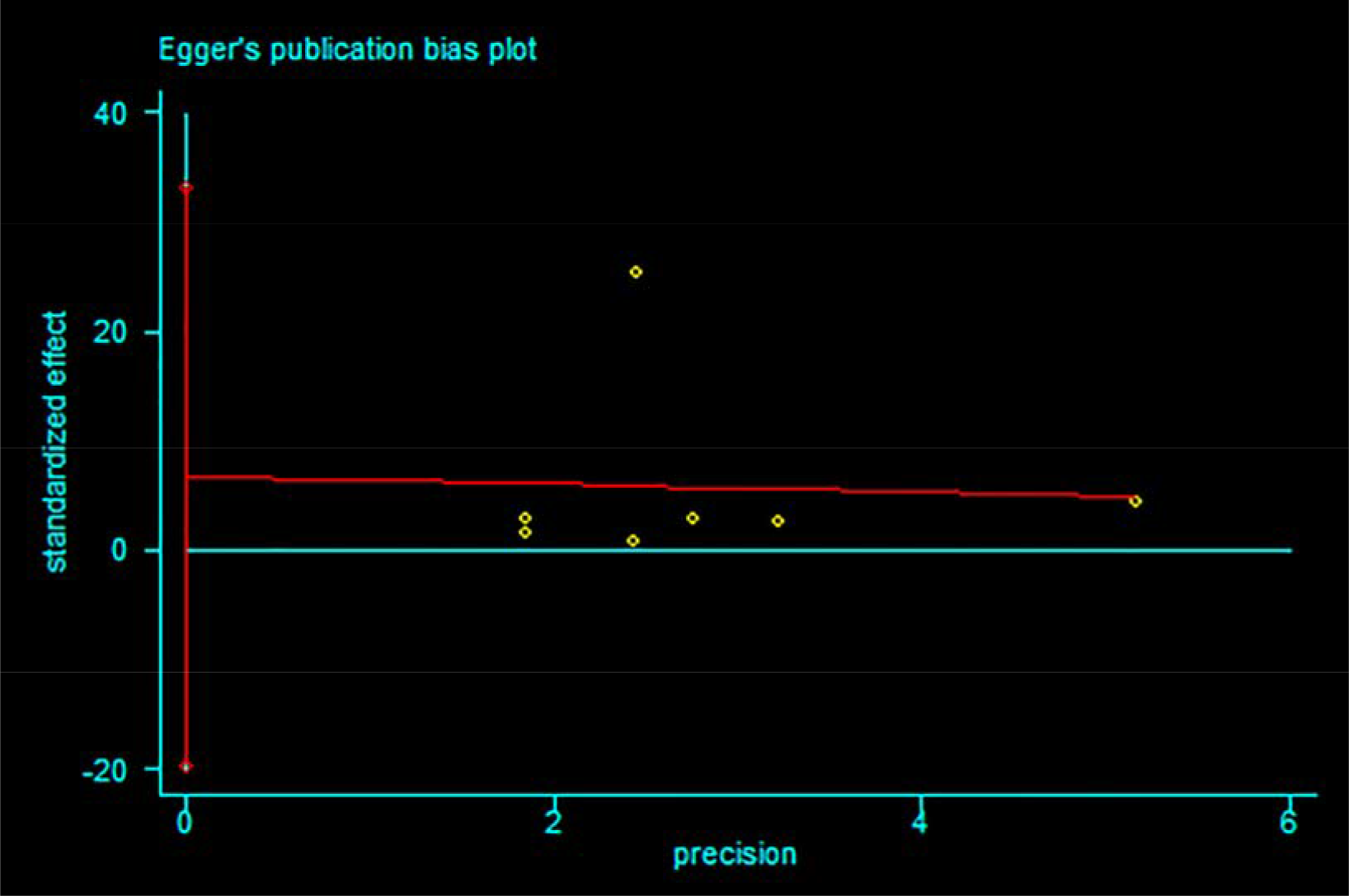
Egger’s funnel plot of TM6SF2 rs58542926 T/C in the uper-dominant model (TT+CC vs. TC,P=0.540).

**Table S1.** Subgroup analysis on the genetic association among TM6SF2 E167K polymorphism and significant liver fibrosis

| Group | Fibrosis |  |  |  |
| --- | --- | --- | --- | --- |
|  | Data points<br>(n) | Pooled OR (95% CI) | P1 | P2 |
| Median age(yr) |  |  |  |  |
| ≤50 | 2 | 1.183 ( 0.656 , 2.131 ) | 0.601 |  |
| >50 | 3 | 1.351 ( 0.197 , 9.248 ) | 0.000 | 0.000 |
| Sample size(n) |  |  |  |  |
| ≤200 | 4 | 2.494 ( 0.522 , 11.903 ) | 0.000 |  |
| >200 | 3 | 0.730 ( 0.472 , 1.130 ) | 0.097 | 0.000 |

NOTE: OR represents the risk of significant fibrosis for TT + CC carriage compared to CT group; P1 represents the heterogeneity of pooled result within each subgroup; P2 represents the intersubgroup heterogeneity across studies categorized by the same criteria;

## Notes

### Competing Interest Statement

The authors have declared no competing interest.

### Funding Statement

There was no funding support.

### Author Declarations

There is nothing suitable

